# Plasma brain-derived tau and phosphorylated tau at T181 are elevated in amyotrophic lateral sclerosis

**DOI:** 10.1101/2025.06.26.25330363

**Authors:** Tiziana Petrozziello, Evan Mizerak, Aparna Krishnamoorthy, Rachel A. Donahue, Ayleen L. Castillo Torres, Ranee Zara B. Monsanto, Bruno L. Hammerschlag, Hannah A. Webster, Becky Fillingham, Pia Kivisäkk, Jamie Timmons, Kelly Fox, Steven E. Arnold, Joshua Cohen, Justin Klee, Sabrina Paganoni, Merit E. Cudkowicz, Lori B. Chibnik, James D. Berry, Ghazaleh Sadri-Vakili

## Abstract

There is an unmet need for reliable biomarkers for amyotrophic lateral sclerosis (ALS). Recent studies demonstrated that the levels of the microtubule-associated protein tau are altered in plasma and cerebrospinal fluid (CSF) from people living with ALS. Our previous findings revealed that while the ratio between tau and phosphorylated tau at T181 (pTau-T181) is lower in ALS CSF compared to healthy controls (HC), higher CSF tau levels correlated with faster disease progression. Here, we measured total tau and pTau-T181 in plasma samples from two cohorts of participants with ALS and HC using two platforms: Quanterix Simoa and Meso Scale Discovery (MSD). Both assays demonstrated significantly higher pTau-T181 and pTau-T181:tau ratio levels in ALS compared to HC. Longitudinal analysis revealed that higher pTau-T181 levels at baseline correlated with faster decline on the revised ALS functional rating scale (ALSFRS-R). Although the MSD analysis revealed higher plasma tau levels in ALS, the Quanterix Simoa assay demonstrated lower total tau levels in ALS plasma than in HC. To investigate the source of this discrepancy, we measured brain-derived tau (BD-tau) using a new Quanterix assay that specifically detects tau originating from the central nervous system (CNS). This analysis demonstrated significantly elevated plasma BD-tau levels in ALS, mirroring the results obtained with the MSD assay, suggesting that the elevated plasma tau observed in ALS is primarily CNS-derived. Collectively, our data indicate that plasma BD-tau and pTau-T181 levels are elevated in ALS, and future studies will aim to define their potential utility as diagnostic or prognostic biomarkers.

## Introduction

Amyotrophic lateral sclerosis (ALS) is a progressive and devastating neurodegenerative disease characterized by a marked heterogeneity in both clinical manifestations and underlying pathogenic mechanisms.^1–3^ The ALS clinical presentation differs by age of onset, region of onset and progression rate and pattern.^1^ This variability complicates clinical diagnosis and ALS therapy development.^4^ Therefore, reliable diagnostic and/or prognostic biomarkers for ALS could hasten diagnosis, aid in clinical prognosticating and help improve ALS trial design.

Recent studies have begun to link alteration in the microtubule-associated protein tau (MAPT) to ALS.^5–11^ For example, several studies have demonstrated cytoplasmic inclusion of tau phosphorylated at several epitopes, including T175, S208/210, S212, T217, S396 and S404, in ALS post-mortem brain and spinal cord tissues,^5,6,8,9,11^ as well as in post-mortem brains from individuals with ALS/frontotemporal dementia (ALS/FTD)^7^ and individuals with concomitant chronic traumatic encephalopathy (CTE) and ALS (CTE-ALS).^10^ Furthermore, previous studies have shown that tau and phosphorylated tau (pTau) are altered in both CSF and plasma derived from individuals with ALS.^5,12–24^ Similarly, our previous study demonstrated that CSF total tau levels were significantly elevated in people with bulbar onset ALS,^5^ and more importantly, CSF total tau levels correlated with faster disease progression.^5,20,22,24^ Furthermore, our study was one of six reports that reliably recapitulated the finding that CSF pTau-T181:tau ratio is lower in ALS participants,^5,17–19,24^ and positively correlated with the score on the revised ALS functional rating scale (ALSFRS-R).^5^ Recent findings have also demonstrated that there is an increase in plasma pTau-T181 in ALS,^12–16^ suggesting that alterations in total tau and pTau-T181 levels in ALS biofluids could potentially serve as a disease monitoring, prognostic or diagnostic biomarker of ALS. Having said that, each of these biomarkers has different characteristics, correlation with disease onset and progression and will be of benefit in contexts of use. Building on previously published findings, we sought to determine whether plasma total tau, pTau-T181 and pTau-T181:tau ratio are altered in ALS to help further clarify their potential use as biomarkers. Here we used two separate cohorts of samples and two methods, namely Quanterix Simoa and Meso Scale Discovery (MSD) assays, to assess total tau and pTau levels.

## Materials and Methods

### Human plasma samples

A random sampling of de-identified ALS and healthy control (HC) plasma samples and accompanying clinical information were obtained from the Northeast Amyotrophic Lateral Sclerosis Consortium (NEALS) Biofluid Repository at Mass General Hospital. The samples were collected and submitted to the biorepository from participants in multi-site research studies between 2008 and 2023. Blood was collected using K2EDTA tubes, and centrifuged at 1,750 g for 10 min. The supernatants were then collected, aliquoted into cryovials and frozen at −80°C within 2 h of collection. Sample were stored below −70°C, with no freeze-thaw cycles.

This study involved participants grouped in two large and independent cohorts from the NEALS repository but with no overlapping participants, defined as ‘cohort 1’ and ‘cohort 2’.

Cohort 1 included longitudinally collected samples from HC (n = 52) and individuals with ALS (n = 76). HC group included 50% male individuals, while ALS group included 63% male individuals. Mean age at screening for HC was 48.4 years (SD = 16.1), while mean age for ALS individuals was 61.8 years (SD = 11.2). Thirteen participants with ALS (22%) had bulbar onset disease, while 40 individuals (68%) had limb onset ALS.

Cohort 2 included samples collected from HC (n = 90) and individuals with ALS (n = 98) only from a single time point. HC group from cohort 2 included 54% male individuals, while ALS group included 51% male individuals. Mean age at screening for HC was 45.7 years (SD = 7.8) while mean age for ALS individuals was 45.8 years (SD = 8.3). Ten individuals with ALS (13%) had bulbar onset disease, while 66 individuals (84%) had limb onset ALS.

Individuals with ALS from cohorts 1 and 2 had similar baseline characteristics, except that participants from cohort 2 were younger and had shorter disease duration (median disease duration = 14.0 months, (Q1, Q3) = (7.7, 22.8)) compared to individuals from cohort 1 (median disease duration = 35.51 months, (Q1, Q3) = (16.0, 47.0)). The average ALSFRS-R score was lower in participants with ALS from cohort 1 (mean ALSFRS-R score = 35, SD = 7.9) compared to individuals with ALS from cohort 2 (mean ALSFRS-R score = 38.3, SD = 6.9). Demographics of participants with ALS and HC from both cohorts are reported in Table 1.

**Table 1:** Baseline characteristics for ALS cohorts 1 and 2.

|  | Cohort 1 |  | Cohort 2 |  |
| --- | --- | --- | --- | --- |
|  | HC (n = 52) | ALS (n = 76) | HC (n = 90) | ALS (n = 98) |
| Age at sample collection, mean (SD), n | 48.4 (16.1), n=36 | 61.8 (11.2), n=59 | 45.65 (7.77), n=76 | 45.78 (8.30), n=96 |
| Sex, n male (%), n | 18 (50%), n=36 | 37 (63%), n=59 | 41 (54%), n=76 | 49 (51%), n=96 |
| pTau-T181, median [Q1, Q3], n | 14.36 [2.24, 20.70], n=41 | 24.73 [6.86, 43.94], n=76 | 2.01 [1.56, 2.54], n=85 | 5.19 [2.86, 8.41], n=96 |
| Total Tau, median [Q1, Q3], n | 1.40 [0.97, 2.00], n=48 | 0.72 [0.54, 1.03], n=76 | 22.23 [17.98, 29.61], n=90 | 27.92 [21.12, 42.92], n=98 |
| pTau-T181:total tau Ratio, median [Q1, Q3], n | 8.67 [1.25, 21.72], n=36 | 28.64 [6.63, 71.76], n=76 | 0.09 [0.07, 0.13], n=85 | 0.15 [0.09, 0.24], n=96 |
| BD-tau, median [Q1, Q3], n | 5.58 [4.61, 6.52], n=28 | 8.70 [6.72, 12.17], n=31 |  |  |
| **ALS disease characteristics** |  |  |  |  |
| Age at symptom onset, mean (SD), n |  | 58.9 (11.5), n=59 |  | 43.5 (8.9), n=96 |
| Site of Onset, Bulbar, n (%) |  | 13 (22%), n=59 |  | 10 (13%), n=79 |
| Site of Onset, Limb, n (%) |  | 40 (68%), n=59 |  | 66 (84%), n=79 |
| Disease Duration, Median [Q1, Q3], n |  | 25.0 [16.0, 47.0], n=59 |  | 14.0 [7.7, 22.8], n=89 |
| ALSFRS-R, mean (SD), n |  | 35.0 (7.9), n=47 |  | 38.3 (6.9), n=81 |
| Pre-baseline ALSFRS, mean (SD), n |  | 0.58 (0.80), n=47 |  | 1.42 (2.16), n=77 |

### Quanterix Simoa assay

Plasma levels of pTau-T181, total tau and BD-tau were measured using Simoa Assay kits (#104111, pTau-181 Advantage V2.1 Kit; #101552, Tau Advantage Kit; #104797, BD-Tau Advantage PLUS Kit, Quanterix Corporation, Billerica, MA) on a fully automated Quanterix HD-X Analyzer at the Mass General Institute for Neurodegenerative Disease (MIND) Biomarker Core at Massachusetts General Hospital according to manufacturer’s instructions and as previously reported.^5,25^ Briefly, plasma samples were centrifuged at 10,000 g for 5 min, diluted 1:4 in Quanterix sample diluent, and measured in duplicate across two runs for total tau and BD-tau, and three runs for pTau-T181. 3-6 internal analytical controls spanning the range of the analytes were included in all runs for pTau-T181, total tau, and BD-tau to account for run-to-run variability. The functional LLOQ was 8 pg/mL for pTau-T181, 0.06 pg/mL for total tau, and 0.533 pg/mL for BD-tau. The coefficient of variance (CV) for duplicates was 9.5%±8.1% for total tau, 6.4%±6.2% (mean±SD) for pTau-T181, and 4.1%±4.3% for BD-tau. The standardized means of the analytical controls across the runs fell within the predefined acceptance limit of 80-120% of the mean for pTau-T181 and BD-tau but not for total tau (68-132%). No run-to-run normalization was consequently performed for pTau-T181 and BD-tau, while total tau was normalized using the analytical controls.

### Meso Scale Discovery assay

Plasma tau and pTau-T181 were measured by using a ECLIA S-Plex Human Tau (total) kit (#K151APSS, Meso Scale Discovery), and a S-Plex Human Tau (pT181) kit (#K151AGMS, Meso Scale Discovery) according to manufacturer’s instructions. Briefly, samples were centrifuged at 2,000 g for 3 min after thawing. Undiluted samples, calibrators and QCs were added to coated plates and incubated for 90 min at room temperature (RT). Next, the plates were incubated consecutively with the turbo-boost, enhancer and turbo-tag (detection) solutions. Lastly, MSD read buffer was added before analysis in an MSD reader.

### Statistics

Normal distribution of measured data was not assumed regardless of sample size or characteristics. The data are presented as bar graphs demonstrating individual values with the whiskers representing the standard error or as noted individual value plots with the central line representing the median and the whiskers representing the interquartile range, as noted. For plasma analysis, the pre-baseline ALSFRS-R slope was calculated as the [(48 – baseline ALSFRS-R score)/(months between onset of ALS weakness or spasticity and the baseline study visit)]. When longitudinal data is available, summary slopes of ALSFRS-R total score and the plasma measures were estimated from the observed values using a mixed effects model with a fixed effect for time and a random intercept and slope for each participant with an unstructured covariance. Additionally, participant-level slopes in plasma measures and ALSFRS-R, measured as unit change per month, were compared using linear regression with ALSFRS-R slope as outcome and each plasma measure modeled separately. Adjustment for disease duration was considered as a confounder. For cross-sectional data, comparisons between groups were performed using a non-parametric Mann-Whitney *U* test and correlations of plasma measures with clinical measures were assessed using non-parametric Spearman correlations. Receiver operating characteristics (ROC) curves were plotted to compare discriminatory performance of individual plasma measures and a combination of multiple measures within a cohort. Area under the ROC curve (AUC) and ROCs were calculated using a logistic regression with the outcome of ALS versus HC. To enable model comparison within cohort 1 regarding BD-tau measurements, the ROC curves and AUC were calculated but were limited to individuals with complete data. Comparisons with clinical measures were not corrected for multiple comparisons. All tests were two sided with a significance level of 0.05, and exact *p* values are reported. Analyses were performed using GraphPad Prism 10.6.1 and R, a language and environment for statistical computing (https://www.R-project.org/).

### Study approval

All human samples used in this study were de-identified. The collection, storage and sharing of these samples was approved by the Mass General Brigham Institutional Review Board (Boston, MA, USA).

## Results

### Total tau and pTau-T181 levels are altered in ALS plasma

To determine whether tau phosphorylation could serve as biomarkers for ALS, we measured total tau, pTau-T181 and their ratio in two large and independent cohorts − both from the NEALS biorepository, but with no overlapping participants. Each consisted of plasma samples derived from participants with ALS and HC. These cohorts are referred to as ‘cohort 1’ and ‘cohort 2’ respectively. Baseline characteristics for both cohorts are presented in Table 1.

First, total tau, pTau-T181, and pTau-T181:tau ratio were assessed in cohort 1 by Quanterix Simoa assay. While average total tau levels were significantly lower in ALS plasma compared to HC (0.88 (SD = 0.53) vs 1.49 (SD = 0.61), p < 0.0001) (Figure 1A), pTau-T181 levels in ALS plasma were significantly elevated compared to HC (30.29 (SD = 28.5) vs 12.97 (SD = 10.41), p = 0.0004) (Figure 1B). Similarly, pTau-T181:tau ratio was significantly higher in ALS plasma than in HC (52.07 (SD = 61.25) vs 13.18 (SD = 15.40), p < 0.0001) (Figure 1C).

**Figure 1.**
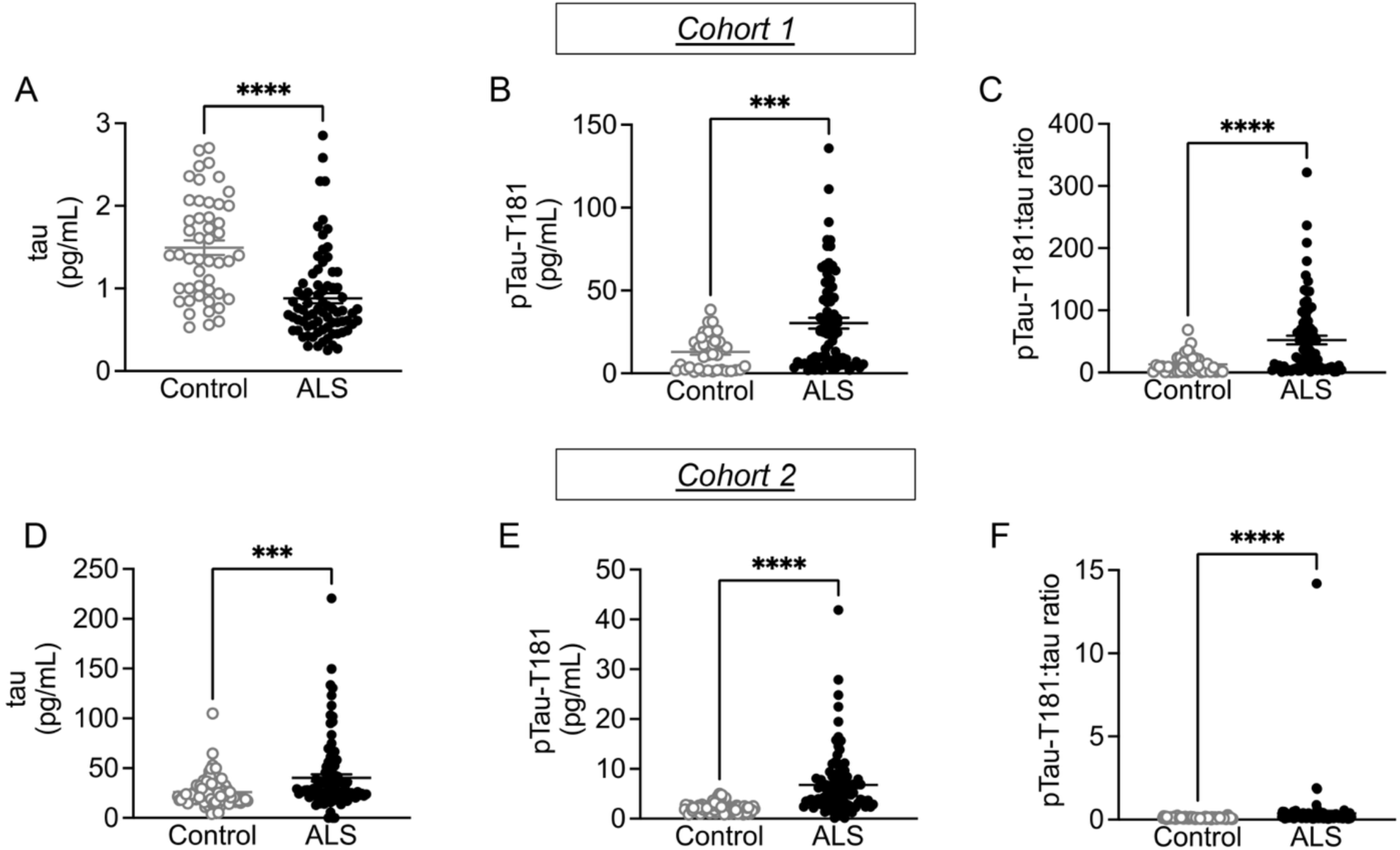
Total tau, pTau-T181 and pTau-T181:tau ratio levels are altered in ALS plasma. While (**A**) tau levels were significantly lower (Mann-Whitney U test = 737.5; p < 0.0001), (**B**) pTau-T181 levels (Mann-Whitney U test = 918.5; p = 0.0004), and (**C**) pTau-T181:tau ratio (Mann-Whitney U test = 701; p < 0.0001) were significantly higher in ALS plasma (n = 76) compared to HC (n = 52) in cohort 1, as measured by Quanterix Simoa Assay. Additionally, (**D**) tau levels (Mann-Whitney U test = 3253; p = 0.0003), (**E**) pTau-T181 levels (Mann-Whitney U test = 1359; p < 0.0001), and (**F**) pTau-T181:tau ratio (Mann-Whitney U test = 2092; p < 0.0001) were significantly higher in ALS plasma (n = 98) compared to HC (n = 90) in cohort 2, as measured by MSD assay. *** p < 0.001; **** p < 0.0001.

Next, we assessed total tau, pTau-T181 levels and their ratio in cohort 2 by MSD assay. Total tau levels in ALS plasma were significantly elevated compared to HC (40.07 (SD = 34.84) vs 25.85 (SD = 13.76), p = 0.0003) (Figure 1D). Similarly, higher pTau-T181 levels were observed in ALS plasma compared to HC (6.77 (SD = 6.27) vs 2.18 (SD = 0.94), p < 0.0001) (Figure 1E). Lastly, average pTau-T181:tau ratio was also significantly higher in ALS plasma compared to HC (0.37 (SD = 1.45) vs 0.098 (SD = 0.045), p < 0.0001) (Figure 1F).

### Alterations in plasma tau, pTau-T181 and pTau-T181:tau ratio were independent of region of onset

To determine whether measuring tau and pTau-T181 levels could help stratify individuals with ALS based on region of disease onset, we correlated the levels with bulbar vs limb onset for participants from whom this information was available.

In cohort 1, total tau levels were lower in both bulbar- (0.82 (SD = 0.48), p = 0.0001) (Figure 2A) and limb-onset ALS (0.80 (SD = 0.57), p < 0.0001) (Figure 2D) compared to HC. pTau-T181 levels were higher in bulbar-onset (29.07 (SD = 19.21), p = 0.0016) (Figure 2B) and limb-onset ALS (28.81 (SD = 29.56), p = 0.0066) (Figure 2E) than in HC. pTau-T181:tau ratio was higher in bulbar- (48.60 (SD = 39.49), p = 0.0001) (Figure 2C) and limb-onset ALS (50.07 (SD = 64.10), p = 0.0004) (Figure 2F) compared to HC.

**Figure 2.**
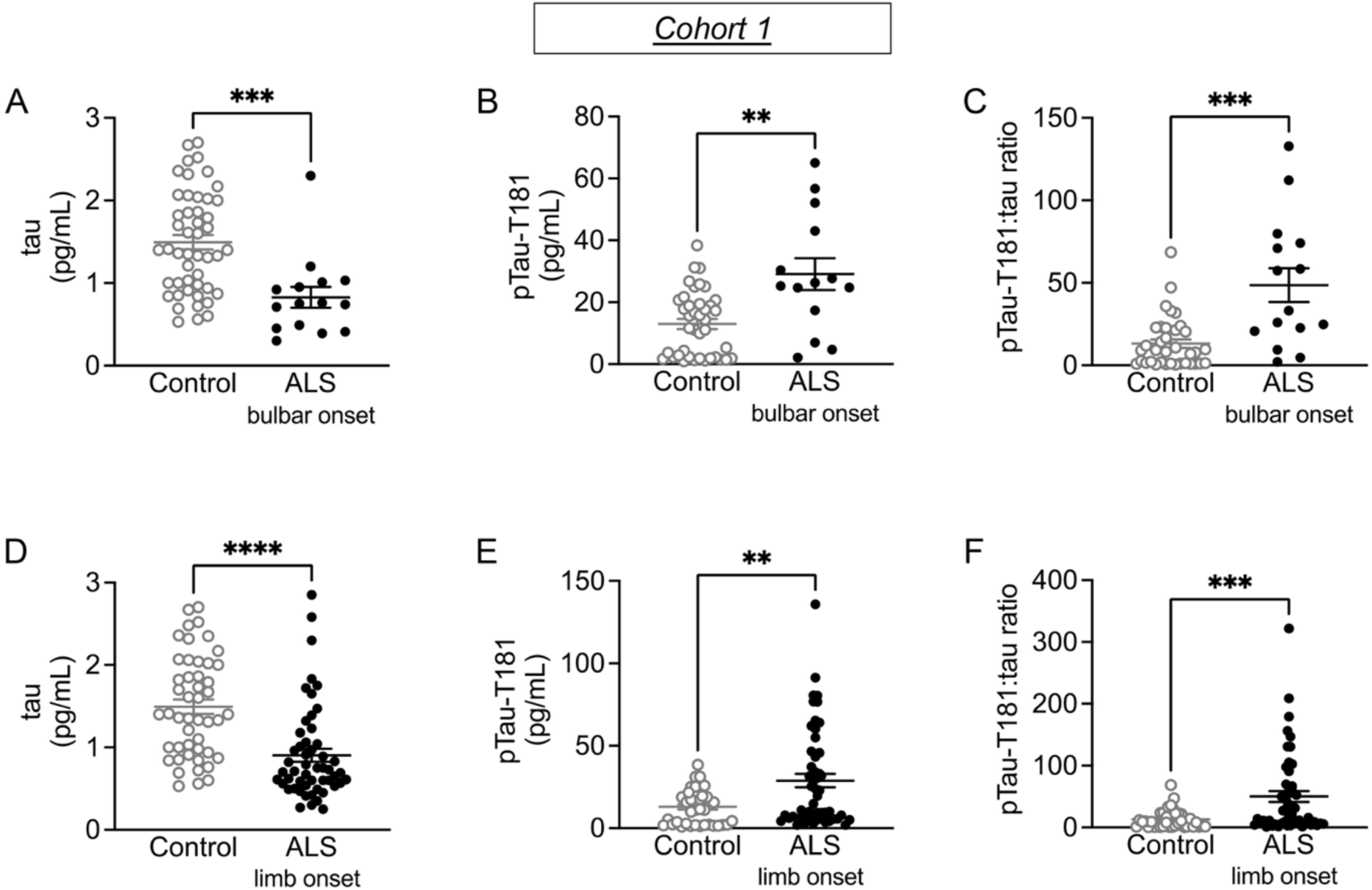
Alterations in tau and pTau-T181 are independent of region of onset in cohort 1. (**A**) Tau levels were significantly lower (Mann-Whitney U test = 131; p = 0.0001), while (**B**) pTau-T181 levels (Mann-Whitney U test = 124; p = 0.0016), and (**C**) pTau-T181: tau ratio (Mann-Whitney U test = 94; p = 0.0001) were significantly higher in bulbar-onset ALS (n = 13) compared to HC (n = 52). Similarly, while (**D**) tau levels were significantly lower (Mann-Whitney U test = 525.5; p < 0.0001), (**E**) pTau-T181 levels (Mann-Whitney U test = 712; p = 0.0066), and (**F**) pTau-T181:tau ratio (Mann-Whitney U test = 526; p = 0.0004) were significantly higher in limb-onset ALS (n = 40) compared to HC (n = 52). ** p < 0.01; *** p < 0.001; **** p < 0.0001.

In cohort 2, total tau levels were higher in both bulbar- (51.24 (SD = 40.59), p = 0.0083) (Figure 3A) and limb-onset ALS (39.50 (SD = 36.26), p = 0.0051) (Figure 3D) compared to HC. pTau-T181 levels were higher in bulbar-onset (8.13 (SD = 5.96), p = 0.0005) (Figure 3B) and limb-onset ALS (6.45 (SD = 5.12), p < 0.0001) (Figure 3E) compared to HC. pTau-T181:tau ratio was not different in bulbar-onset ALS (0.18 (SD = 0.12), p = 0.0600) (Figure 3C), while it was higher in limb-onset ALS (0.43 (SD = 1.73), p < 0.0001) (Figure 3F) compared to HC.

**Figure 3.**
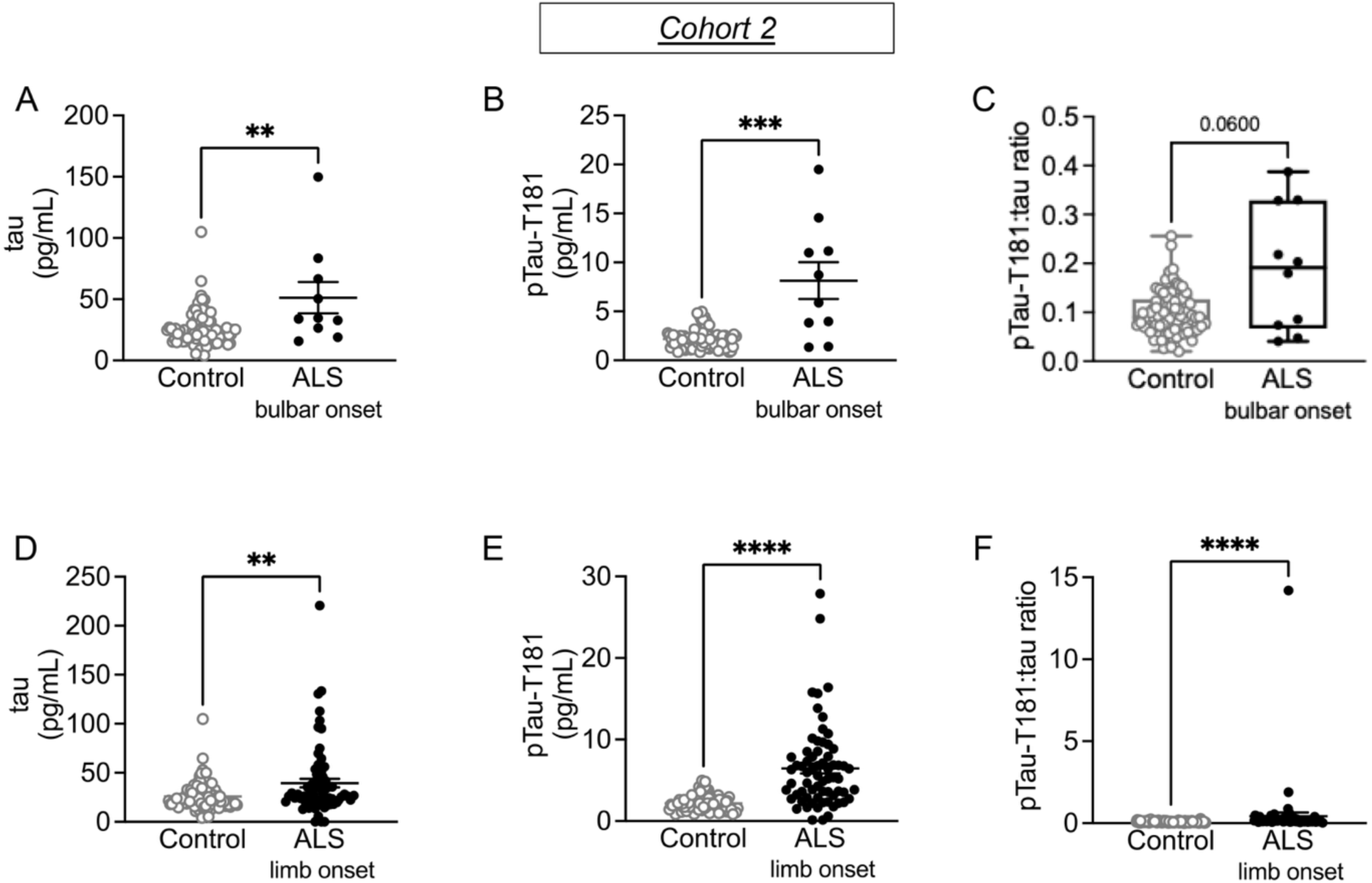
Alterations in tau and pTau-T181 are independent of region of onset in cohort 2. While (**A**) tau levels (Mann-Whitney U test = 235; p = 0.0083), and (**B**) pTau-T181 levels (Mann-Whitney U test = 158; p = 0.0005) were significantly higher, (**C**) pTau-T181: tau ratio (Mann-Whitney U test = 283; p = 0.0600) was not changed in bulbar-onset ALS (n = 10) compared to HC (n = 94). Similarly, (**D**) tau levels (Mann-Whitney U test = 2336; p = 0.0051), (**E**) pTau-T181 levels (Mann-Whitney U test = 880; p < 0.0001), and (**F**) pTau-T181:tau ratio (Mann-Whitney U test = 1324; p < 0.0001) were significantly higher in limb-onset ALS (n = 66) compared to HC (n = 94). ** p < 0.01; *** p < 0.001; **** p < 0.0001.

### Plasma pTau-T181 levels correlated with faster ALS progression

Next, we correlated tau and pTau levels with clinical characteristics, including age, disease duration, and ALSFRS-R at the time of sample collection.

Our results demonstrated no statistically significant correlation between either age or disease duration and total tau, pTau-T181, and pTau-T181:tau ratio at the time of sample collection in cohort 1 (Supplementary Figure 1). Similarly, no statistically significant correlation was found between either age or disease duration and total tau, pTau-T181 and pTau-T181:tau ratio levels in cohort 2 (Supplementary Figure 2).

In cohort 1, ALSFRS-R at baseline as well as pre-baseline ALSFRS-R slope had no correlation with total tau, pTau-T181 and pTau-T181:tau ratio at the time of sample collection (Supplementary Figure 3); similarly, in cohort 2, both ALSFRS-R at baseline and pre-baseline ALSFRS-R slope had no correlation with total tau, pTau-T181 and pTau-T181:tau ratio at the time of sample collection (Supplementary Figure 4).

Longitudinal analysis of ALS plasma from cohort 1 demonstrated a variability in the trajectories of tau, pTau-T181, and pTau-T181:tau ratio levels (Table 2). As expected, ALSFRS-R scores declined at an average rate of 0.48 (SD = 0.05) points per month (p < 0.001). Total tau revealed a statistically significant increase of 0.02 pg/mL (SD = 0.01) per month (p = 0.007), whereas pTau-T181 showed an average increase of 0.15 pg/mL (SD = 0.14) per month increase (p = 0.27) and pTau-T181:tau ratio showed an average decrease of 0.10 pg/mL (SD = 0.36) decrease per month (p = 0.79) although not statistically significant (Table 2). Importantly, higher pTau-T181 levels correlated with faster decline in ALSFRS-R over time, with a 1 pg/mL higher pTau-T181 level associated with a 0.20 points per month faster slope in ALSFRS-R decline (beta (SE) = 0.20 (0.07), 95%CI (0.06, 0.34), p = 0.007). Whereas total tau demonstrated a positive, yet non-significant association with ALSFRS-R decline (Beta (SE) = 1.18(1.69), 95%CI (−2.22, 4.59), p = 0.487), pTau-T181:tau ratio revealed no association (Beta(SE) = 0.00 (0.02), 95%CI (−0.03, 0.03), p = 0.960).

**Table 2.** Trajectories of ALSFRS-R, total tau, pTau-T181, and pTau-T181: tau ratio in ALS plasma from cohort 1.

|  | **Slope** | **Slope SE** | **p-value** |
| --- | --- | --- | --- |
| ALSFRS-R | -0.48 | 0.05 | < 0.001 |
| tau | 0.02 | 0.01 | 0.007 |
| pTau-T181 | 0.15 | 0.14 | 0.273 |
| pTau-T181:tau ratio | -0.1 | 0.36 | 0.787 |

\* Slope is measured as change per month, p-value tests if slope is different from 0

### BD-tau levels were elevated in ALS plasma

Given that our findings revealed opposite directions in total tau levels in ALS plasma in two large and independent cohorts as compared to HC (lower in cohort 1 vs higher in cohort 2) and as measured by two different assays (Quanterix Simoa vs MSD assays), we sought to clarify the source of this discrepancy. To this aim, we used the novel BD-tau assay developed by Quanterix which recognizes low molecular weight isoforms of tau lacking the large peptide in exon 4a found in peripheral tissues.^26^ Specifically, we measured BD-tau levels in a subset of ALS (n = 31) and HC (n = 28) participants from cohort 1.

Our results demonstrated that BD-tau levels in ALS plasma were significantly higher than in HC (10.07 (SD = 5.36) vs 5.65 (SD = 1.60), p < 0.0001) (Figure 4A).

**Figure 4.**
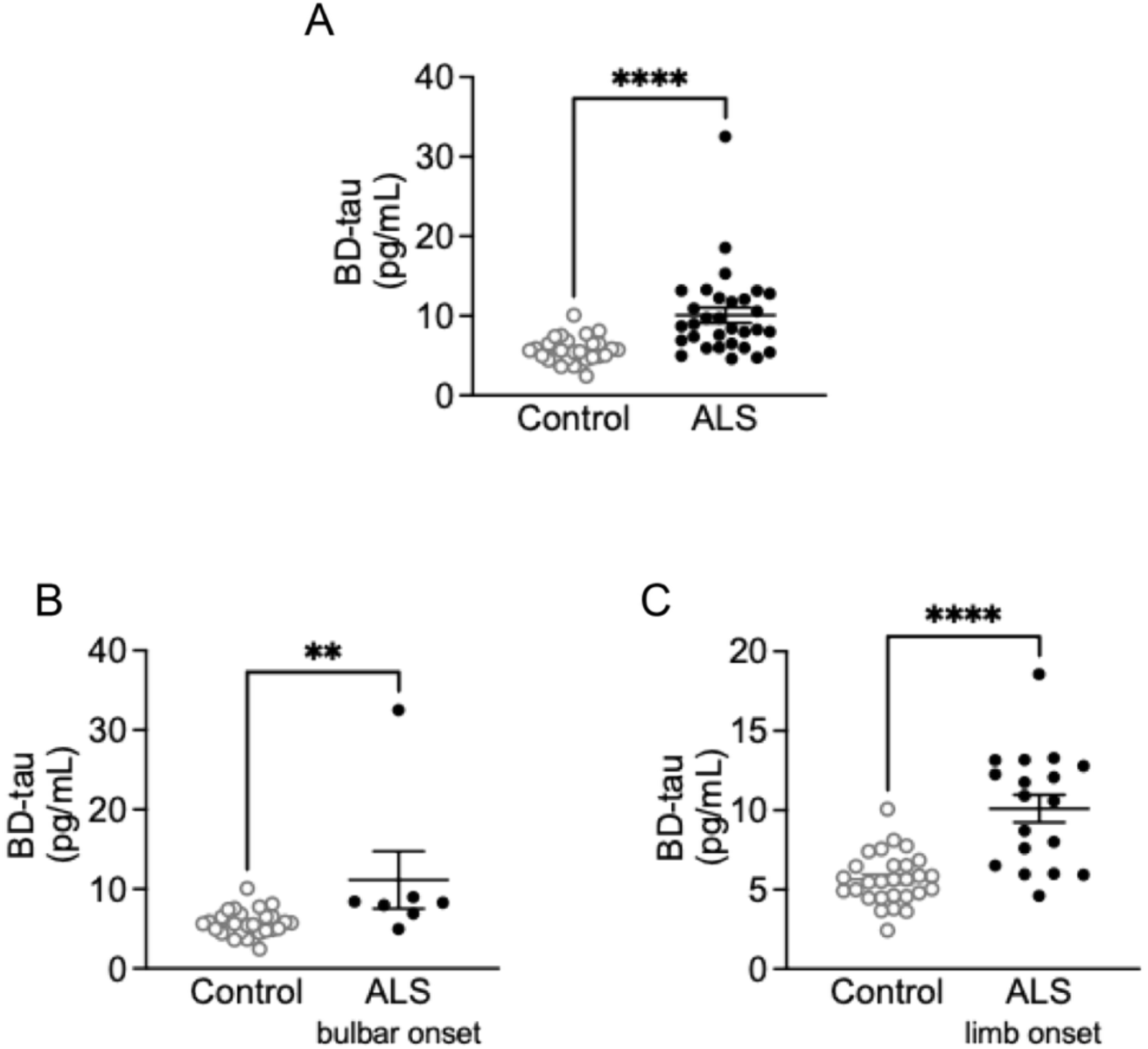
BD-tau levels were significantly elevated independently of region of onset in a subset of ALS plasma from cohort 1. (**A**) BD-tau levels were significantly higher in ALS (n = 31) than in HC (n = 28) (Mann-Whitney U test = 132; p < 0.0001). (**B**) BD-tau levels were significantly higher in bulbar-onset ALS individuals (n = 7) compared to HC (n = 28) (Mann-Whitney U test = 28; p = 0.0025). (**C**) BD-tau levels were significantly higher in limb-onset ALS participants (n = 18) than in HC (n = 28) (Mann-Whitney U test = 61; p < 0.0001).

Similar to total tau, we sought to determine whether measuring BD-tau could help stratify individuals with ALS based on region of disease onset. Therefore, in participants with known information on region of ALS onset, we correlated BD-tau levels with bulbar vs limb onset.

BD-tau levels were significantly higher in both bulbar- (11.15 (SD = 9.51), p = 0.0035) (Figure 4B) and limb-onset ALS (10.10 (SD = 3.65), p < 0.0001) (Figure 4C) compared to HC.

Lastly, we correlated BD-tau levels with clinical characteristics, including age, disease duration, and ALSFRS-R at the time of sample collection.

Our results demonstrated no statistically significant correlation between either age or disease duration and BD-tau at the time of sample collection (Supplementary Figure 3A and B). Similar, both ALSFRS-R at baseline and pre-baseline ALSFRS-R slope had no correlation with BD-tau at the time of sample collection (Supplementary Figure 3C and D).

### Plasma tau measures discriminate between ALS and HC

To assess the discriminatory performance of each plasma measure (total tau, pTau-T181, pTau-T181:tau ratio, and BD-tau), we generated ROC curves and calculated the corresponding AUC values.

In cohort 1, when analyzed individually, the AUC were 0.784 (95% CI: 0.696, 0.872) for total tau, 0.718 (95% CI: 0.620, 0.816) for pTau-T181, and 0.744 (95% CI: 0.651, 0.837) for pTau-T181:tau ratio. Of note, a model including both total tau and pTau-T181 yielded the highest AUC at 0.791 (95% CI: 0.706, 0.876) (Figure 5A).

**Figure 5.**
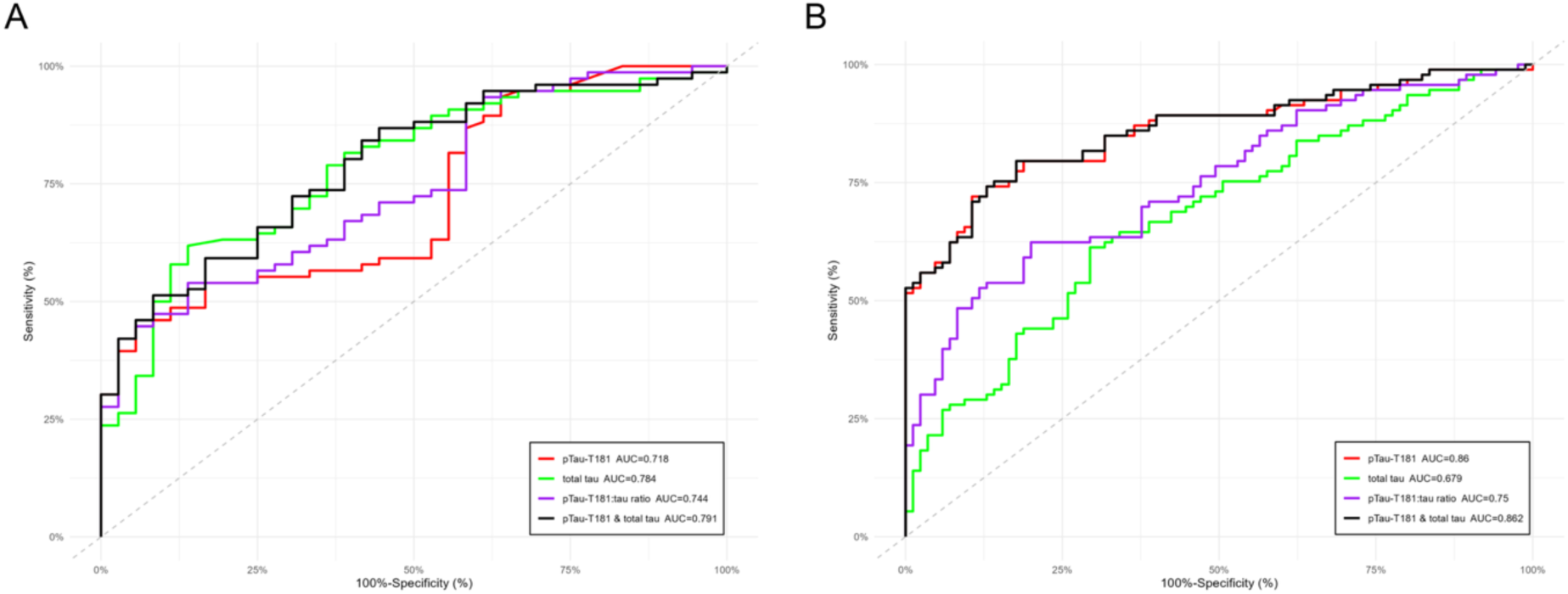
Plasma measures discriminate between ALS and HC in cohorts 1 and 2. (A) Comparison of individual plasma measures (total tau, pTau-T181, and pTau-T181:tau ratio) in cohort 1 (n = 112). The graph also includes a model combining total tau and pTau-T181. (B) Comparison of individual plasma measures (total tau, pTau-T181, and pTau-T181:tau ratio) in cohort 2 (n = 181). The graph also includes a model combining total tau and pTau-T181.

In cohort 2, when analyzed individually, the AUC were 0.679 (95% CI: 0.601, 0.757) for total tau, 0.860 (95% CI: 0.805, 0.915) for pTau-T181, and 0.750 (95% CI: 0.679, 0.820) for pTau-T181:tau ratio (Figure 5B). Similar to cohort 1, the combined model again demonstrated the best performance at 0.862 (95% Cl: 0.807, 0.916) (Figure 5B).

Because BD-tau was measured only in a subset of cohort 1, to properly compare across measures, we re-calculated the AUC in cohort 1 limited to samples that also had a measure for BD-tau. The AUC for all four plasma measures ranged from 0.828 (95% Cl: 0.713, 0.942) for BD-tau to 0.885 (95% Cl: 0.793, 0.978) for pTau-T181. Lastly, the model including both total tau and pTau-T181 again reveled the highest AUC of 0.896 (95% Cl: 0.803, 0.989) (Figure 6).

**Figure 6.**
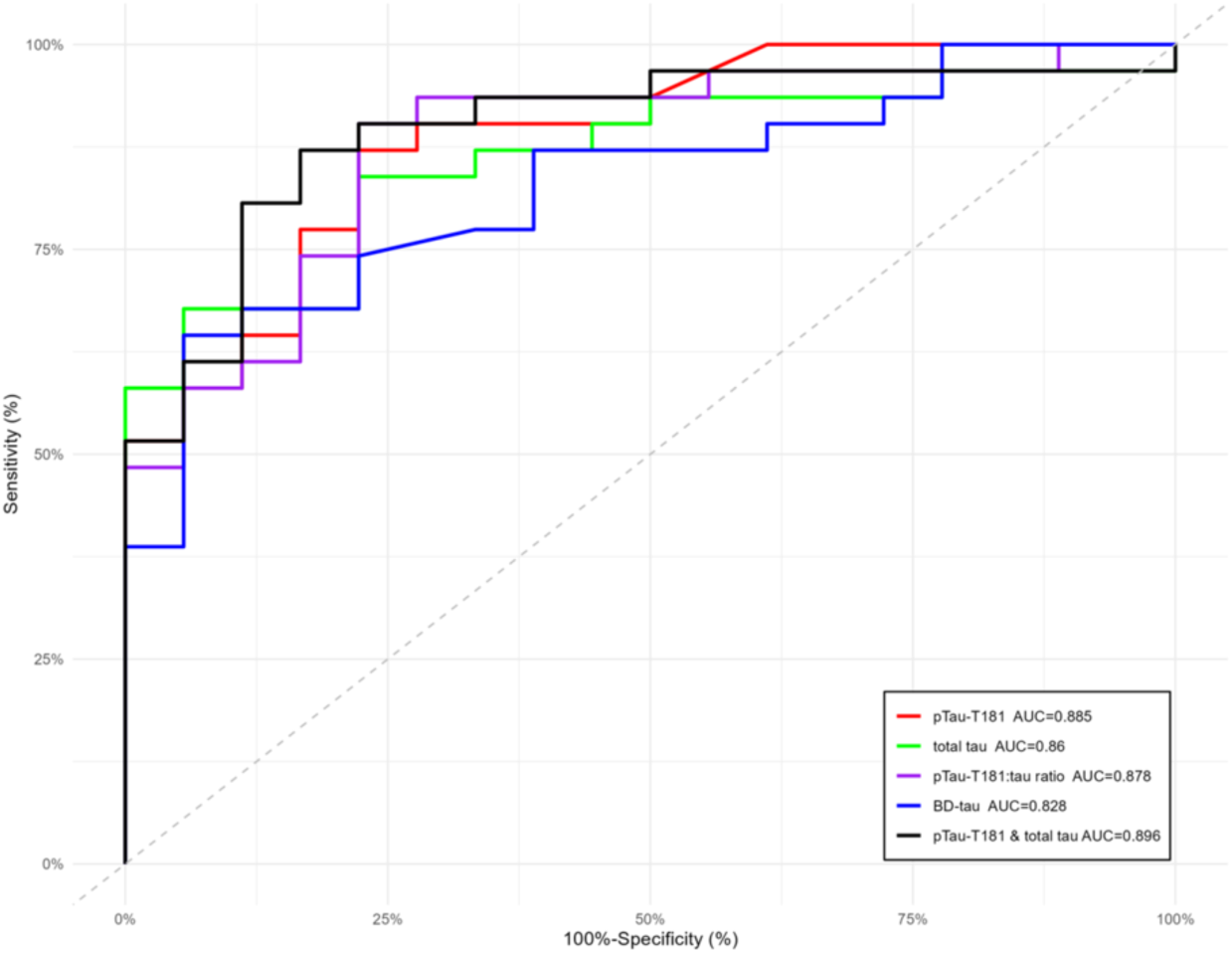
ROC and AUC analysis including BD-tau in cohort 1. Comparison of individual plasma measures, including total tau, pTau-T181, pTau-T181:tau ratio, and BD-tau in cohort 1 (n = 49). The graph also includes a model combining total tau and pTau-T181.

## Discussion

In this study, we found significantly higher levels of plasma total tau (MSD assay and Quanterix BD-tau), pTau-T181, and pTau-T181:tau ratio in ALS compared to HC using two different cohorts of plasma samples. Both assays also revealed that alterations in total tau, pTau-T181 and the ratio occurred with both bulbar and limb-onset ALS. Additionally, longitudinal analyses revealed that elevated plasma pTau-T181 levels correlated with faster decline on the ALSFRS-R, supporting previous findings which revealed elevated plasma pTau-T181 over time in individuals with ALS with faster progressing disease.^12^

Previous studies have suggested that elevations in CSF total tau correlated with faster ALS progression.^5,20,22,24^ However, little has been known about total tau levels in ALS plasma. To our knowledge, only a single study demonstrated lower serum total tau levels in ALS.^27^ Somewhat surprisingly, our results demonstrated lower total tau levels in plasma samples from cohort 1 (Quanterix Simoa assay), but elevated levels in cohort 2 (MSD assay). The lower levels of total tau in ALS compared to HC in cohort 1 was surprising, given that tau levels increase with age,^28,29^ and that the samples in cohort 1 consisted of older individuals with a shorter disease course at time of clinic visit when the samples were collected (Table 1). This discrepancy could be due differences in the cohorts, perhaps from cofounders such as concomitant medications, coexisting FTD, or other unknown differences between the cohorts. Additionally, these results suggest that increases in plasma tau are unlikely to be driven by aging and support a mechanistic rather than age-dependent contribution of tau in ALS given that higher tau levels were found in individuals with shorter ALS duration in cohort 2, and longitudinal analysis in cohort 1 revealed a statistically significant increase in tau levels over time with disease progression (p = 0.007; Table 2). Furthermore, cohort 2 samples consisted only of a single timepoint, and future studies need to include larger plasma cohorts with longitudinal samples that will be tested using both platforms.

Alternatively, the discrepancy in tau levels may reflect differences in assay performance. Although both methods are highly sensitive, their antibodies likely target distinct tau epitopes, resulting in detection of different tau species and thus opposing results. Consistent with this, we observed a significant elevation in BD-tau in a subset of ALS plasma samples from cohort 1. The novel Quanterix BD-tau assay selectively detects brain-derived, low-molecular-weight tau isoforms lacking the exon 4a peptide present in peripheral tau.^26^ These findings suggest that elevated plasma tau in ALS primarily originates from CNS-derived isoforms rather than peripheral ones. Further validation in larger ALS and control cohorts will be necessary to confirm this conclusion.

As for pTau-T181, our findings replicate recently published results demonstrating a significant increase in plasma levels in ALS.^12–16^ Additionally, we also reported a significant higher pTau-T181:tau ratio in ALS plasma compared to HC. Longitudinal analysis in cohort 1 revealed that increases in pTau-T181 correlated with faster disease progression, supporting previous findings,^12^ and suggest that pTau-T181 may be useful as a prognostic biomarker for ALS. It is now clear that CSF pTau-T181 and pTau-T181:tau ratio are lower in ALS,^5,20,22,24^ and do not mirror the increase in plasma as shown here and previously.^12–16^ This discrepancy in CSF vs plasma pTau-T181 levels has a number of plausible explanations. The originating source for plasma may be peripheral tissue as was reported in a recent study demonstrating an increase in pTau-T181 and pTau-T217 in ALS atrophic muscle fibers.^12^ It is also possible that plasma tau is bound to proteins but not in the CSF (matrix effect), or it may be cleared or metabolized quickly in the plasma but not in CSF. Determining which factors are at play will require future studies. Although we focused on pTau-T181 in this study, pTau-T217 and pTau-T231 were shown to be elevated in plasma,^13^ and pTau-T217 was also reported to be increased in ALS serum,^12^ and therefore, these other phospho sites will need to be assessed further.

It should also be noted that, based on our ROC and AUC analyses, the combination of pTau-T181 and total tau demonstrated the strongest discriminatory performance in both cohorts. However, these results should be interpreted in context that the comparison was limited to individuals with ALS versus HC. One of the major caveats of this study is the lack of samples from “ALS mimics.” ALS clinical presentation, especially in the early stages of the disease, overlaps with other neurological disorders including, cerebrovascular disease, cervical myelopathy, neuropathy, and myasthenia gravis to name a few.^30^ Therefore, in order to determine if tau and pTau are viable biomarkers for ALS, future studies will need to include samples from individuals with “ALS mimics” as well as other tauopathies such as AD, to determine whether total tau and pTau-T181 are able to discriminate between other conditions and ALS.

## Data Availability

All data produced in the present study are available upon reasonable request to the corresponding author.

## Ethical Publication Statement

We confirm that we have read the Journal’s position on issues involved in ethical publication and affirm that this report is consistent with those guidelines.

## Data availability statement

The data supporting the findings of this study are available within the article and/or its supplementary material.

## Acknowledgments

The authors would like to thank patients and their families for sample donation.

## Funding statement

T.P. was supported by an award from The Linda and Mike Mussallem Foundation to Massachusetts General Hospital. G.S.V. was supported by the Andre Family. The MIND Biomarker Core was supported by NIA grant P30AG062421.

## Conflict of interest disclosure

SEA reports receiving institutional grant or sponsored research support from the Alzheimer’s Association, Alzheimer Drug Discovery Foundation, Challenger Foundation, John Sperling Foundation, National Institutes of Health, Prion Alliance, AbbVie, Amylyx, Athira Pharma, Eli Lilly, Ionis Pharmaceuticals, Janssen Pharmaceutical/Johnson& Johnson, MarvelBiome, Novartis AG, Seer Bioscience, and SuperFluid; and payments for consulting and/or participation on scientific advisory boards of Allyx Therapeutics, BioVie Pharma, Bob’s Last Marathon, Cassava Sciences, Cortexyme/Quince Therapeutics, EIP Pharma/CervoMed, Foster & Eldredge, Jocasta Neuroscience, Merck, Sage Therapeutics, and Vandria. EM, JT, KF, JC, and JK are full-time employees of and may have stock option ownership in Amylyx Pharmaceuticals, Inc.

## Patient consent statement

Written informed consent was obtained from all participants prior to enrollment in the study.

## Author contributions

T.P. contributed to the study design, data collection, data analysis, and drafting of the manuscript. E.M., A.K., A.L.C.T., R.Z.B.M., B.L.H.M B.F., P.K., J.T., K.F. contributed to the data collection, data analysis, and editing of the manuscript. S.E.A., J.C., J.K., S.P., M.E.C., L.C., J.D.B. contributed to the study design and editing of the manuscript. GSV contributed to the study design, data analysis, drafting of the manuscript.

---

**Supplementary Figure 1.**
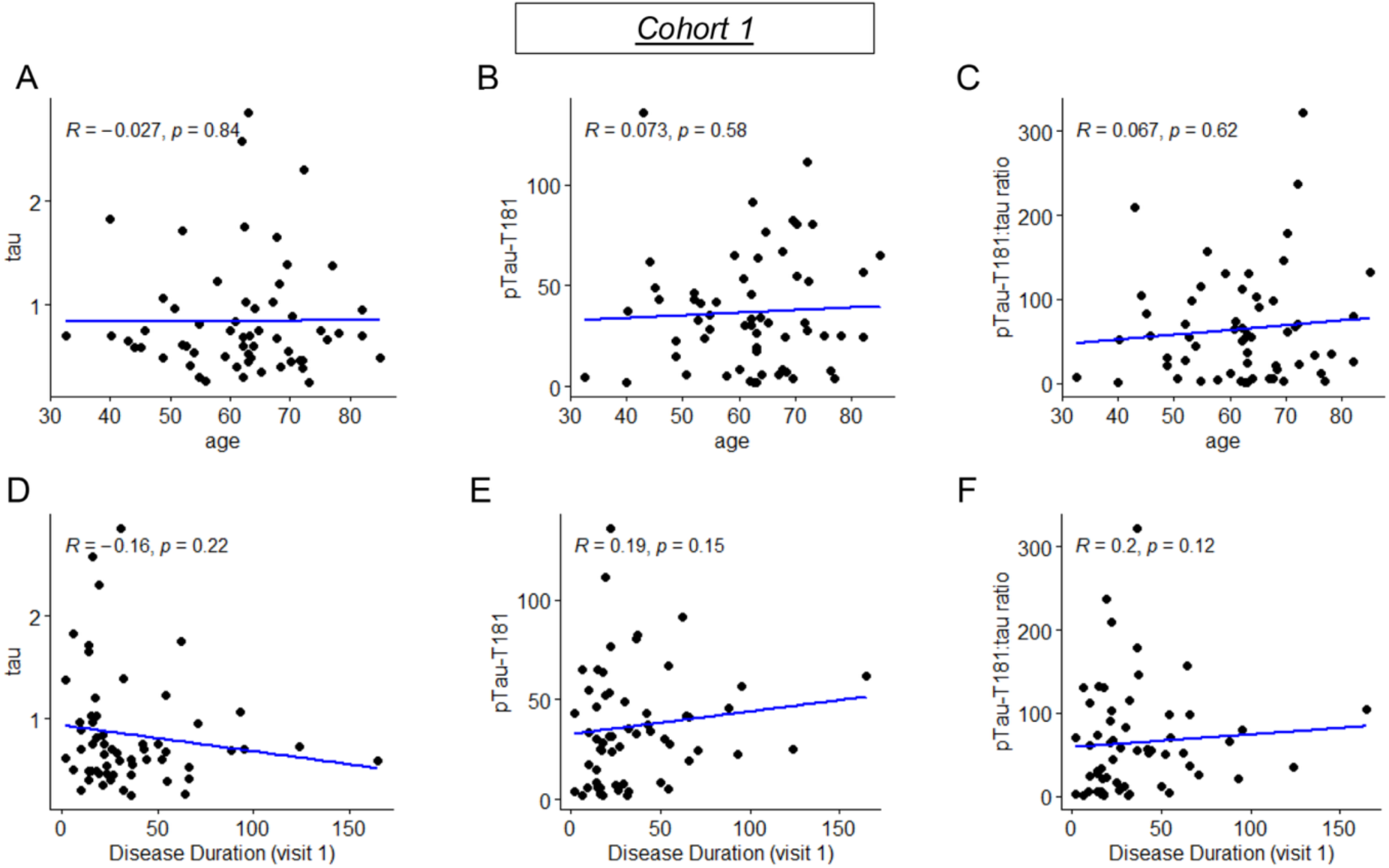
Plasma tau, pTau-T181 and their ratio did not correlate with age and disease duration at the time of sample collection in cohort 1. (**A**) Tau levels (Spearman correlation, R = −0.027; p = 0.84), (**B**) pTau-T181 levels (Spearman correlation, R = 0.073; p = 0.58), and (**C**) pTau-T181:tau ratio (Spearman correlation, R = 0.067; p = 0.62) did not correlate with age at the time of sample collection in cohort 1. (**D**) Tau levels (Spearman correlation, R = −0.16; p = 0.22), (**E**) pTau-T181 levels (Spearman correlation, R = 0.19; p = 0.15), and (**F**) pTau-T181:tau ratio (Spearman correlation, R = 0.2; p = 0.12) did not correlate with ALS duration in cohort 1.

**Supplementary Figure 2.**
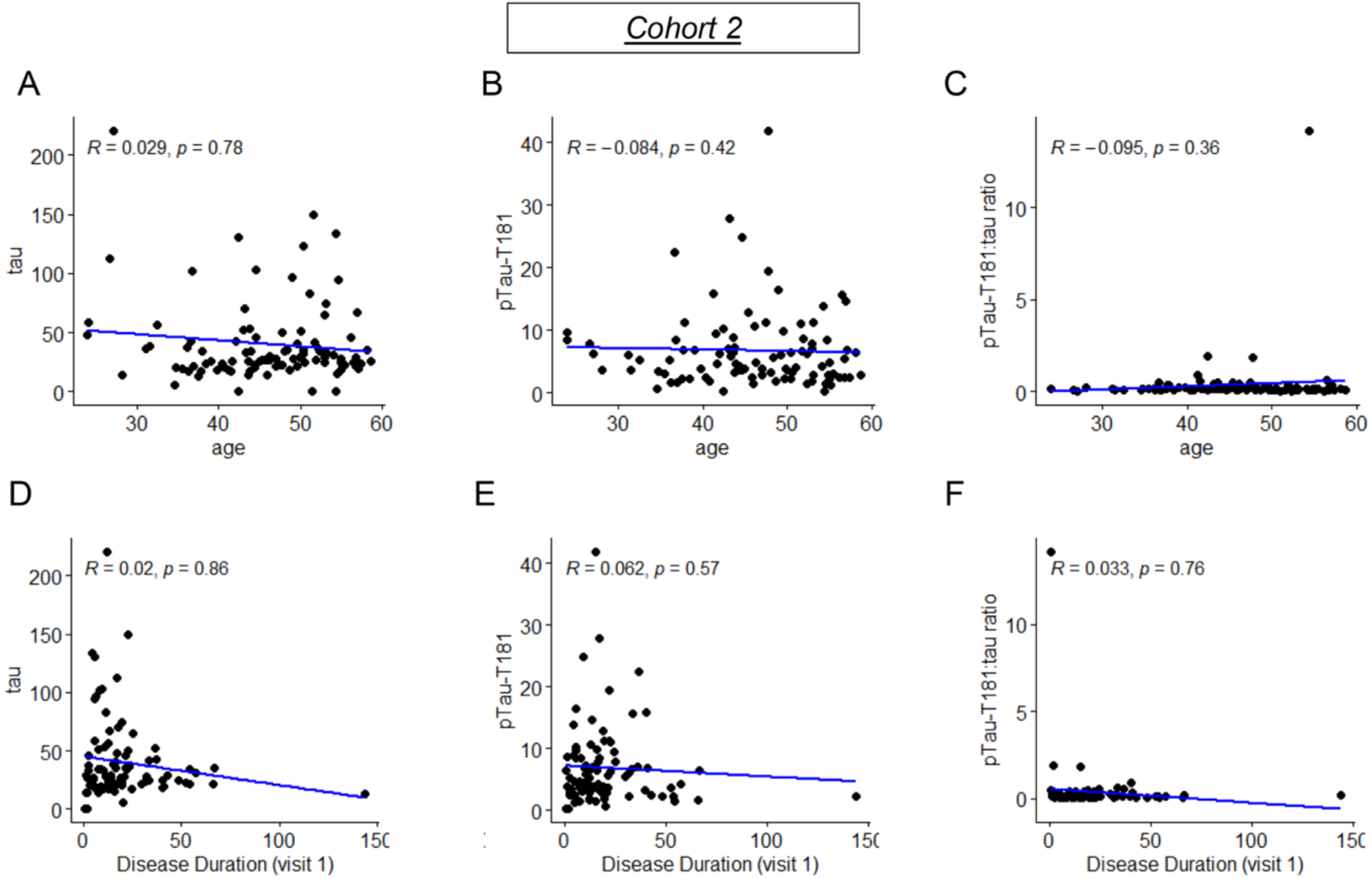
Plasma tau, pTau-T181 and their ratio did not correlate with age and disease duration at the time of sample collection in cohort 2. (**A**) Total tau levels (Spearman correlation, R = 0.029; p = 0.78), (**B**) pTau-T181 levels (Spearman correlation, R = −0.084; p = 0.42), and (**F**) pTau-T181:tau ratio (Spearman correlation, R = −0.084; p = 0.42) did not correlate with age at the time of sample collection in cohort 2. Similarly, (**D**) tau levels (Spearman correlation, R = 0.02; p = 0.86), (**E**) pTau-T181 levels (Spearman correlation, R = 0.062; p = 0.57), and (**F**) pTau-T181:tau ratio (Spearman correlation, R = 0.033; p = 0.76) did not correlate with ALS duration in cohort 2.

**Supplementary Figure 3.**
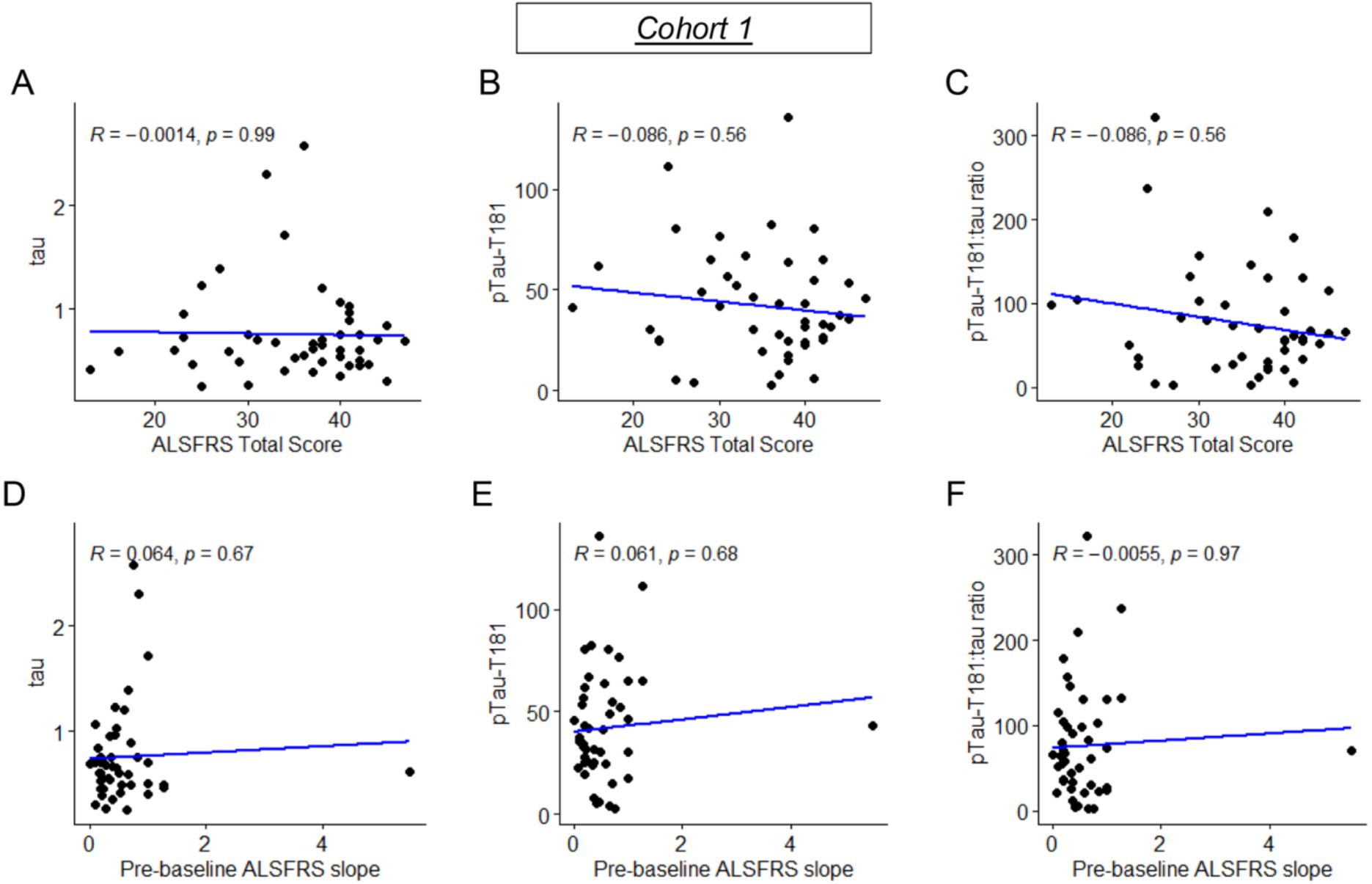
Plasma tau, pTau-T181, and their ratio did not correlate with baseline ALSFRS-R score and pre-baseline ALSFRS-R slope in cohort 1. (**A**) Tau levels (Spearman correlation, R = −0.0014; p = 0.99), (**B**) pTau-T181 levels (Spearman correlation, R = −0.086; p = 0.56), and (**C**) pTau-T181:tau ratio (Spearman correlation, R = −0.086; p = 0.56) did not correlate with the total ALSFRS-R score in cohort 1. Similarly, (**D**) Tau levels (Spearman correlation, R = −0.064; p = 0.67), (**E**) pTau-T181 levels (Spearman correlation, R = −0.061; p = 0.68), and (**F**) pTau-T181:tau ratio (Spearman correlation, R = −0.0055; p = 0.97) did not correlate with the pre-baseline ALSFRS-R slope in cohort 1.

**Supplementary Figure 4.**
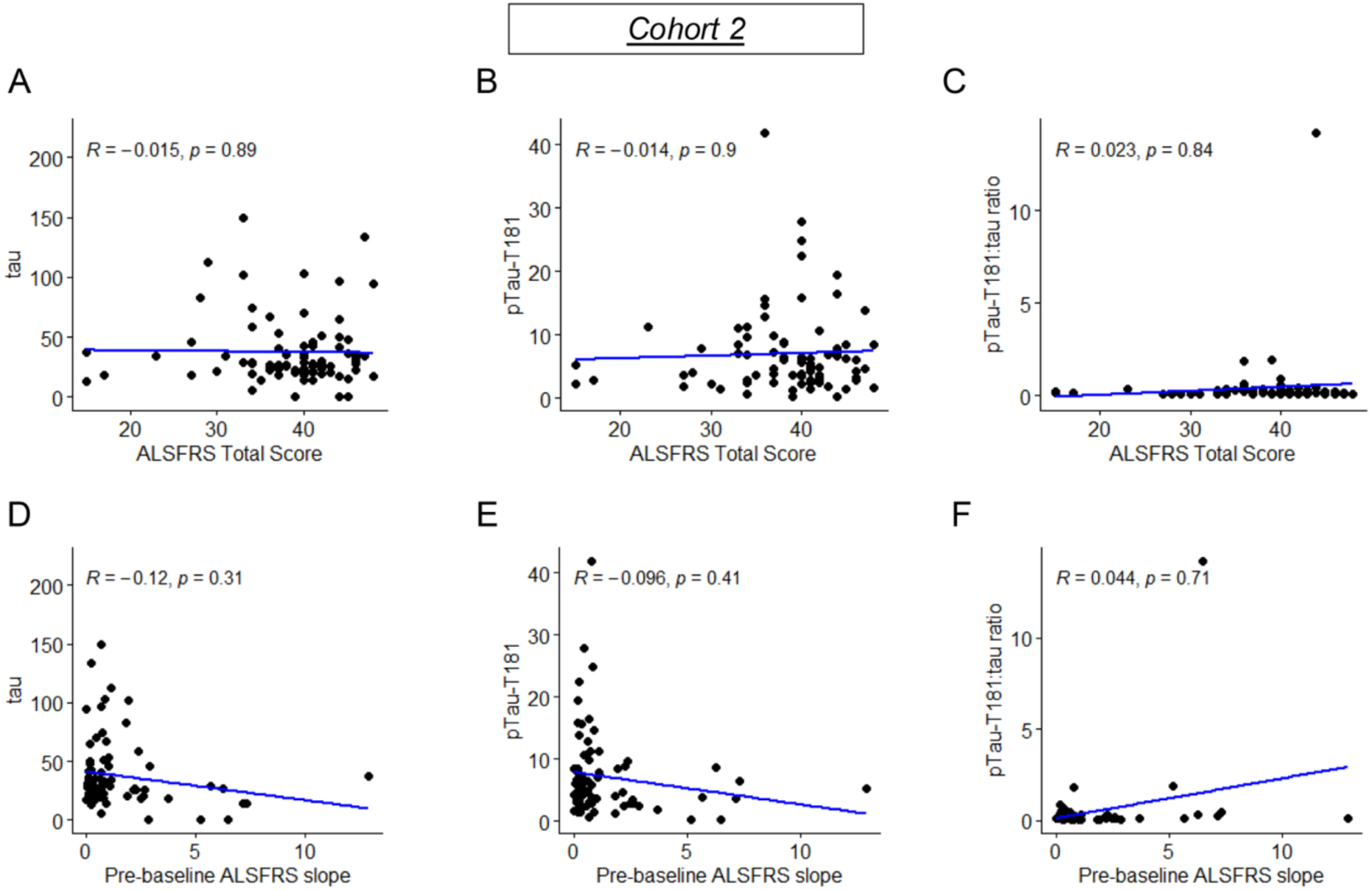
Plasma tau, pTau-T181, and their ratio did not correlate with baseline ALSFRS-R score and pre-baseline ALSFRS-R slope in cohort 1. (**A**) Total tau levels (Spearman correlation, R = −0.015; p = 0.89), (**B**) pTau-T181 levels (Spearman correlation, R = −0.014; p = 0.9), and (**C**) pTau-T181:tau ratio (Spearman correlation, R = 0.023; p = 0.84) did not correlate with the total ALSFRS-R score in cohort 2. Similarly, (**D**) tau levels (Spearman correlation, R = −0.12; p = 0.31), (**E**) pTau-T181 levels (Spearman correlation, R = −0.096; p = 0.41), and (**F**) pTau-T181:tau ratio (Spearman correlation, R = 0.044; p = 0.71) did not correlate with the pre-baseline ALSFRS-R slope in cohort 2.

**Supplementary Figure 5.**
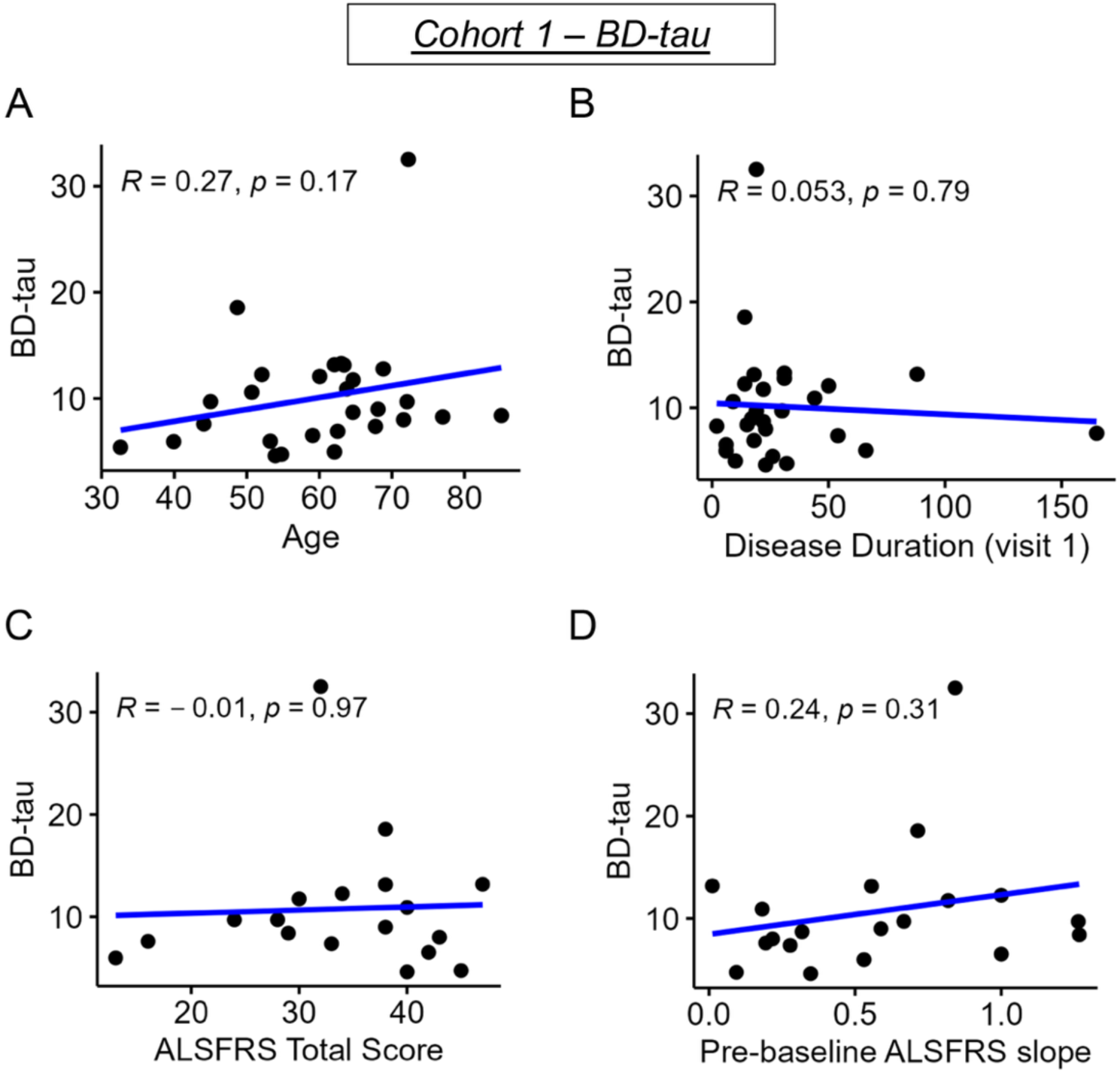
Plasma BD-tau levels did not correlate with age, disease duration, baseline ALSFRS-R score and pre-baseline ALSFRS-R slope in cohort 1. BD-tau levels did not correlate with (**A**) age at the time of sample collection (Spearman correlation, R = 0.27; p = 0.17), (**B**) disease duration (Spearman correlation, R = 0.053; p = 0.79), (**C**) total ALSFRS-R score (Spearman correlation, R = −0.01; p = 0.97), and (**D**) pre-baseline ALSFRS-R slope (Spearman correlation, R = 0.24; p = 0.31) in cohort 1.

## Notes

### Author Declarations

The Mass General Brigham Institutional Review Board (Boston, MA, USA) gave ethical approval for this work.

### Summary of Updates

We have now included new results demonstrating higher brain-derived tau (BD-tau) levels in ALS plasma compared to healthy controls, as measured by the new Quanterix Simoa BD-tau assay.

